# Two Axes of Sleep, One Heart: Dissociating Objective Sleep-Disordered Breathing From Subjective Sleep Duration in Cardiac Risk

**DOI:** 10.64898/2026.11.16.26368255

**Authors:** Qiao Deng, Nai Ma, Taroon Aan, Dongxiao Pang

## Abstract

**Background:** Cardiovascular epidemiology has long treated “sleep” as a single exposure, although it denotes both an objective physiological state and a subjective behaviour. We tested whether these domains carry separable information about cardiac risk.

**Methods and Results:** We derived an objective axis from 8,444 polysomnograms (Sleep Heart Health Study) and a subjective axis of self-reported duration from 407,303 adults in the 2020 and 2022 Behavioral Risk Factor Surveillance System. Multivariable regression (n=213,489), quadratic dose– response modelling, sleep-stratified comorbidity networks, and era-stratified tests were combined. Objective sleep-disordered breathing was continuous and right-skewed (median apnea–hypopnea index, 31.8 events/h; median sleep efficiency, 69%) in a risk-enriched cohort. Both short (<7 h) and long (>9 h) sleep were associated with disease after full adjustment: adjusted odds ratios for heart attack were 1.24 (95% CI, 1.19–1.30) and 1.30 (1.19–1.42), and for stroke 1.12 (1.06–1.17) and 1.42 (1.29–1.57). A positive quadratic term confirmed a U shape for all 4 outcomes formally tested (*P*≤6.3×10⁻²⁰), with fitted nadirs of 7.26 to 7.85 hours. Suboptimal sleep also raised comorbidity-network density from 0.089 to 0.112 (short sleep; +26%) and 0.108 (long sleep; +22%), with non-overlapping bootstrap CIs and concordant effects across demographic strata. A survey-weighted replication in official CDC microdata (n=344,010; median design effect, 4.43) preserved the principal cardiac, U-shaped, and network findings; an exploratory COVID-19 interaction (*P*=0.020) did not survive design-based correction.

**Conclusions:** Objective breathing pathology and subjective duration are separately prognostic, and suboptimal sleep amplifies the cardiometabolic comorbidity network. Duration alone cannot exclude asymptomatic sleep-disordered breathing; cardiovascular prevention should assess both axes.

**Clinical Perspective:** *What Is New?:*

- In 8,444 full-night polysomnograms and 407,303 survey responses, objective sleep-disordered breathing and self-reported sleep duration behave as two separately measurable, non-interchangeable dimensions of sleep exposure.
- Self-reported sleep duration has a formally verified U-shaped relation with cardiometabolic disease, with fitted risk nadirs of 7.26 to 7.85 hours that fall inside the recommended 7–9 hour range.
- Suboptimal sleep is associated with a 22–26% increase in cardiometabolic comorbidity-network density, a structural effect reproduced across every age, sex, and race/ethnicity stratum examined.
- A design-weighted replication in the official CDC microdata (n=344,010; median design effect, 4.43) preserved the principal cardiac, U-shaped, and network associations and identified the secondary findings that are sensitive to the survey design.

*What Are the Clinical Implications?:*

- A reassuring self-report of 7–9 hours of sleep does not exclude severe, asymptomatic sleep-disordered breathing, so patients with hypertension, atrial fibrillation, stroke, or resistant metabolic risk should be considered for objective sleep assessment irrespective of reported duration.
- A report of more than 9 hours of sleep warrants clinical attention as a marker of occult multimorbidity rather than being dismissed as healthy behaviour.
- The Sleep Network Modulation Index offers a compact, reproducible summary of how a modifiable exposure couples multiple disease clusters simultaneously, and may be tracked in longitudinal cohorts or prevention programmes.

**Graphical Abstract:** The Two-Axis Sleep Dissociation Model integrates objective polysomnography (SHHS, n=8,444) with subjective survey sleep (BRFSS, n=407,303). Their intersection defines four cardiac-risk phenotypes; three empirical findings are summarised at right. Quadratic-term *P*≤6.3×10⁻²⁰ refers to the four outcomes formally tested for curvature. All estimates shown are from the unweighted primary analysis; the design-weighted replication is reported in Section 3.8, Table S2, and Figure S1.

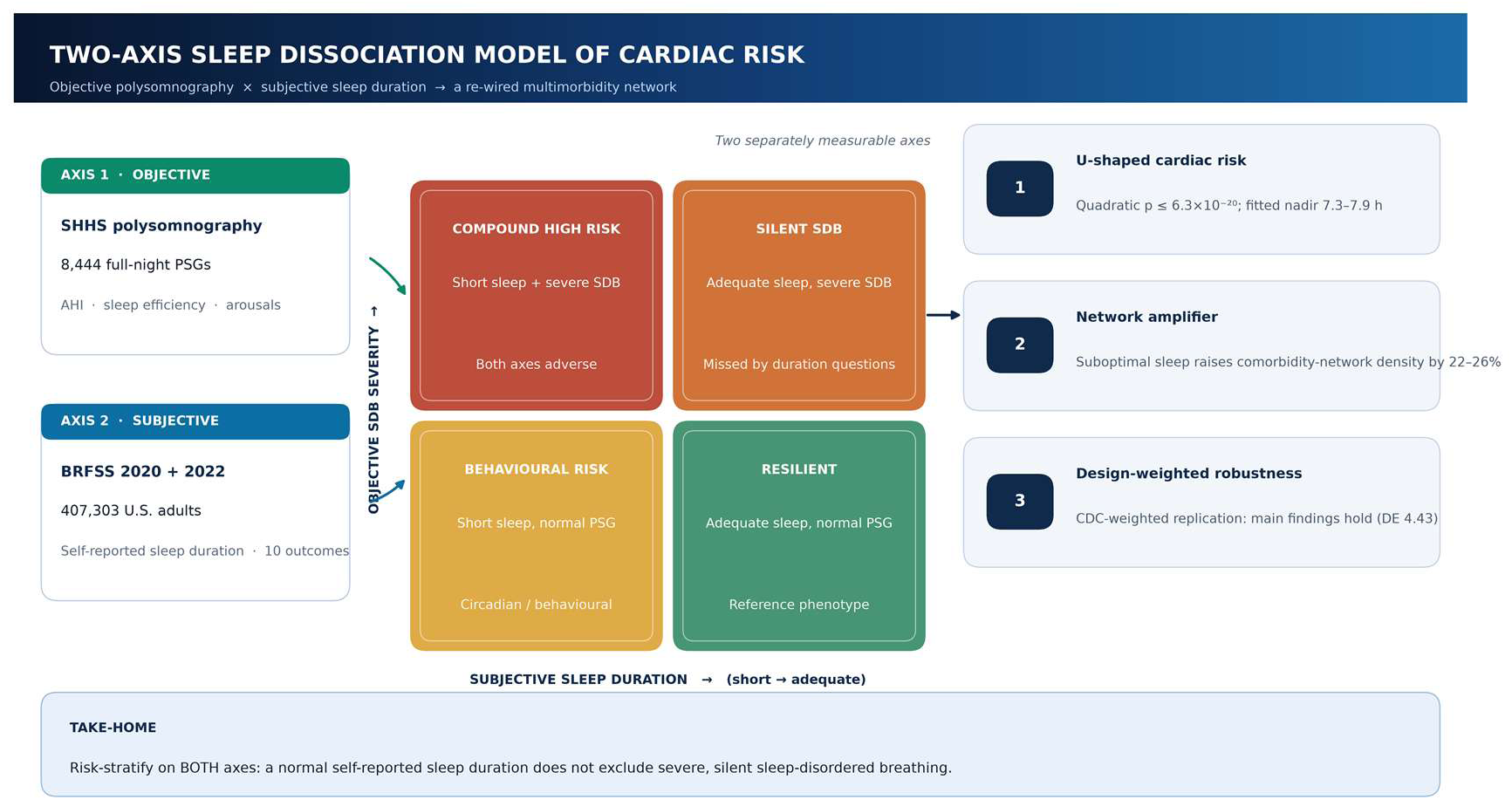

## Introduction

That sleep matters to the heart is no longer in dispute. Three decades of work, anchored by the Sleep Heart Health Study (SHHS), established that objectively measured sleep-disordered breathing (SDB) predicts incident hypertension, stroke, and heart failure, and the American Heart Association has recently summarised the evidence linking obstructive sleep apnoea to cardiovascular disease.^1–4^ In parallel, large prospective cohorts and their meta-analyses described a U-shaped relation between *self-reported* sleep duration and mortality, stroke, and cardiovascular events, with excess risk at both ends of the distribution.5–8 Contemporary frameworks treat sleep as a multidimensional construct rather than a single number,^9^ and habitual short sleep has been linked to adverse cardiovascular health profiles and greater resource use.^10^ Yet these literatures have developed largely in isolation: one records breathing and sleep stages with polysomnography (PSG); the other records a single answer to “how many hours do you sleep?” The implicit, rarely examined assumption is that both are measuring the same exposure.

We argue that they are not. We advance a **Two-Axis Sleep Dissociation Model** (Figure 1), in which cardiac risk reflects two conceptually distinct, separately measurable dimensions:

- **Axis 1 — objective sleep pathophysiology.** Derived here from PSG: the apnea–hypopnea index (AHI), sleep efficiency, stage architecture, and cortical arousals. It captures an anatomical, intermittent-hypoxia and sleep-fragmentation burden that is frequently *asymptomatic*.
- **Axis 2 — subjective sleep behaviour.** Derived here from survey: self-reported duration and its deviation from the 7–9 hour range. It captures circadian, behavioural, and socioeconomic exposure.

**Figure 1.**
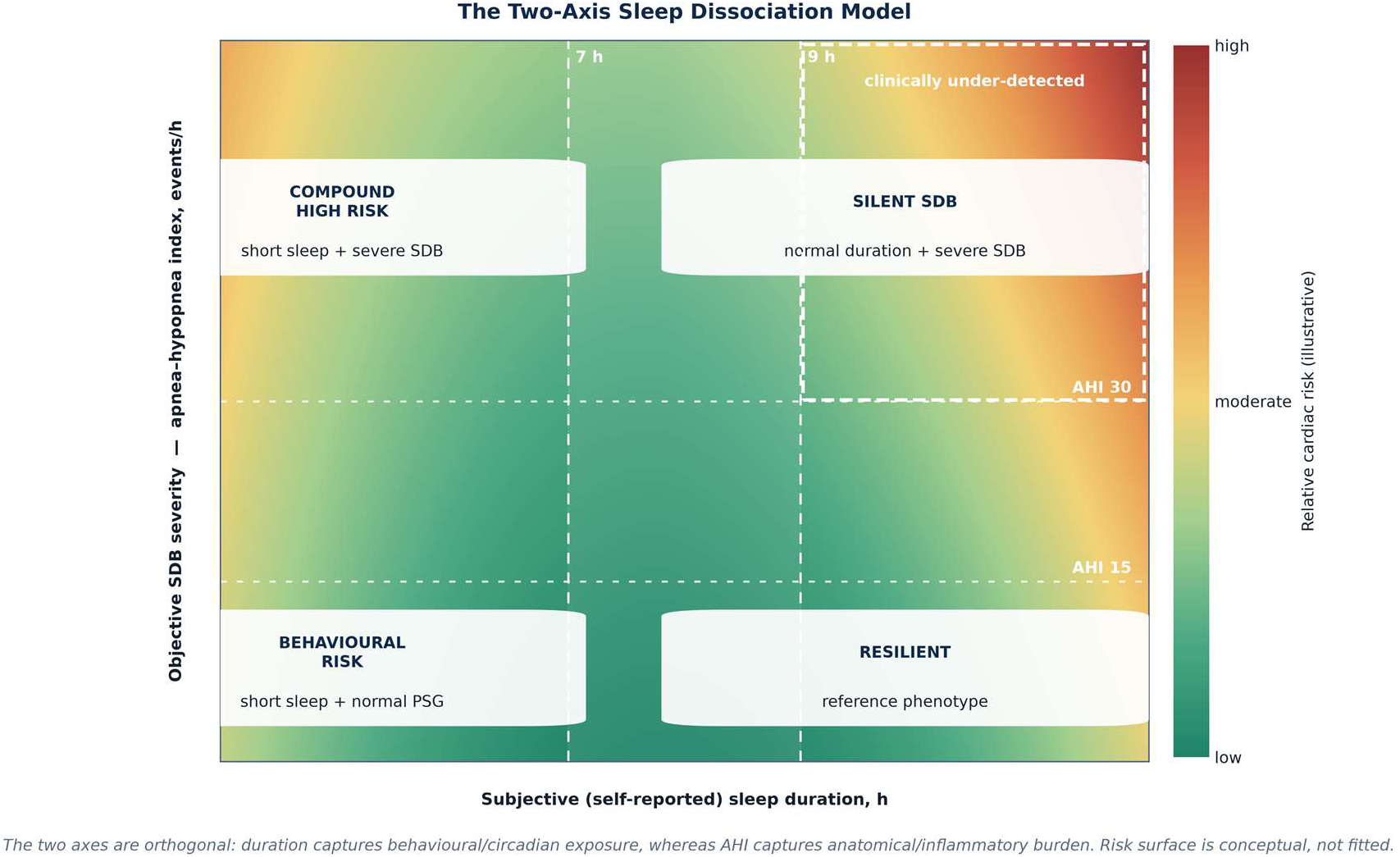
Conceptual Two-Axis model. The horizontal axis is self-reported sleep duration and the vertical axis objective SDB severity (AHI). Four quadrants correspond to resilient, behavioural-risk, silent-SDB, and compound-high-risk phenotypes. The risk surface is illustrative (not fitted); dashed lines mark 7 hours, 9 hours, and AHI 15/30 thresholds. The silent-SDB quadrant is highlighted as clinically under-detected.

The distinction matters clinically. A patient may report perfectly adequate sleep while harbouring severe, undiagnosed obstructive sleep apnoea — the *silent SDB* quadrant of Figure 1 — or may sleep few hours with an entirely normal PSG (*behavioural risk*). Because the two axes rest on different mechanisms, and because SDB is substantially underdiagnosed in the community,^11^ a normal answer on Axis 2 cannot be taken as reassurance about Axis 1. Asking only “do you sleep enough?” therefore risks overlooking a sizeable high-risk group.

Two further questions, neither of which has to our knowledge been addressed at scale, motivated this work. **First**, does sleep duration merely move the *level* of cardiovascular risk, or does it reorganise the *structure* of multimorbidity? Network medicine has shown that diseases cluster in non-random ways, that these clusters can be represented as phenotype networks, and that the choice of co-occurrence measure materially affects the resulting network.^12–15^ Sleep has not, however, been modelled as a modulator of the cardiometabolic comorbidity network. **Second**, because the coronavirus disease 2019 (COVID-19) pandemic superimposed a systemic inflammatory and behavioural stressor on the same population, did the sleep–cardiac association change over that interval, and does prior infection modify it?

Bringing together 8,444 PSG records and 407,303 survey responses, we test the Two-Axis framework and report four findings: the objective axis is continuous and severely distributed within a high-risk PSG cohort; the subjective axis exhibits a formally verified U shape with nadirs inside the guideline range; suboptimal sleep acts as a *network amplifier* of multimorbidity; and the sleep–cardiac association intensified during the pandemic era, with outcome-specific effect modification by prior infection.

## Methods

### 2.1 Study design, reporting, and data provenance

This is a secondary analysis of two independent, de-identified, cross-sectional data sources. Objective sleep phenotypes were characterised in SHHS; subjective sleep, outcomes, and effect modification were analysed in BRFSS. The two cohorts do not overlap at the individual level and were therefore **not pooled**; each informs a different axis of the model. Reporting follows the Strengthening the Reporting of Observational Studies in Epidemiology (STROBE) recommendations for cross-sectional studies where applicable.^16^

#### Ethics

The analysis was deemed exempt by the the institutional review board of Ningbo No. 2 Hospital, and that written informed consent was not required because only de-identified, publicly distributed research data were used. The parent studies obtained their own approvals and consents. Access to NSRR/SHHS resources is governed by the National Sleep Research Resource data-use terms; BRFSS microdata are distributed by the US Centers for Disease Control and Prevention.

#### Independent data access and analysis

Dr Deng had full access to all the data in the study and takes responsibility for its integrity and the data analysis.

#### Survey design

The primary analyses use cleaned derivative BRFSS files that do not retain the survey’s complex-design variables; they are therefore **unweighted complete-case analyses**, and their prevalences and odds ratios describe the analytic sample rather than design-weighted population quantities. To quantify the consequences of that constraint, we obtained the official CDC LLCP 2020 and 2022 microdata, which do contain the final weight (_LLCPWT), stratification variable (_STSTR), and primary sampling unit (_PSU), and repeated every principal analysis with design-based estimation (Section 2.5, Results 3.8, Table S2, Figure S1). Because the derivative files carry no respondent identifier, the weighted analysis is an **independent replication on the official microdata** — with weighted and unweighted estimates computed on identical records — rather than a reweighting of the primary records. Unless explicitly labelled as design-weighted, all estimates in the main text are unweighted.

### 2.2 Data sources

#### Objective axis — Sleep Heart Health Study

The NSRR-standard PSG annotation archive (shhs.zip) contained 8,444 full-night ScoredEvents XML files (5,793 SHHS-1; 2,651 SHHS-2). Each file encodes sleep stages (Wake, N1–N3, REM), respiratory events (obstructive, mixed and central apnoea; hypopnoea), arousals, and oxygen desaturations, each with a start time and duration. SHHS is a community-based cohort that deliberately oversampled participants at elevated risk of SDB^1,2^; it is used here to characterise the *shape and structure* of Axis 1, not to estimate population prevalence.

#### Subjective axis and outcomes — BRFSS

We analysed heart_2020_cleaned (161,280 respondents) and heart_2022_no_nans (246,023 respondents), giving 407,303 records in total. The 2022 file supplies self-reported SleepHours, 10 binary cardiometabolic and mental-health conditions (heart attack, angina, stroke, chronic obstructive pulmonary disease, diabetes, kidney disease, depressive disorder, arthritis, asthma, skin cancer), prior COVID-19 positivity, vaccination variables, and covariates (age, sex, body mass index, smoking, alcohol, physical activity, general health, and race/ethnicity). The 2020 file supplies analogous fields with SleepTime, a composite HeartDisease item, and Stroke. A duplicate Chinese-language header row present in the source CSV files was removed before analysis.

#### Design-weighted replication source

We additionally read the CDC-distributed SAS transport files LLCP2020.XPT (401,958 records) and LLCP2022.XPT (445,132 records). Following CDC’s complex-sampling guidance for core-component analyses, design-based models used _LLCPWT as the final weight, _STSTR as the stratification variable, and _PSU as the PSU; because _PSU is the annual sequence number and is unique to each record, every respondent constitutes one PSU (equivalent to id = ∼1 in R survey). Sleep duration (SLEPTIM1), the nine chronic-condition items, and all covariates were recoded to the definitions used in the primary analysis. Coding concordance was verified against the derivative files (for example, 2022 heart-attack prevalence 5.68% versus 5.46%, depressive disorder 20.67% versus 20.58%), and the unweighted replication reproduced the main-text aORs to within 0.03 (Table S2, Panel F). Skin cancer is not included in the 2022 core, so the weighted network comprises nine conditions.

### 2.3 Deriving the objective sleep axis

Each PSG XML was parsed for ScoredEvent elements. We computed total sleep time (TST = N1 + N2 + N3 + REM, excluding Wake), time in bed (TIB, from the “Recording Start Time” event), sleep efficiency (TST/TIB), stage proportions, AHI ([apnoea + hypopnoea count]/TST-hours), and arousal index (arousals/TST-hours). Records with TST ≤0 were excluded; all 8,444 valid records were retained. AHI severity was classified using conventional bands (<5, 5–<15, 15–<30, ≥30 events/h).

### 2.4 Defining the subjective sleep axis and outcomes

SleepHours/SleepTime were restricted to the physiologically plausible interval 4–12 hours, excluding approximately 0.4% of records with implausible values. Three groups were defined a priori: **short** (<7 h), **adequate** (7–9 h; reference), and **long** (>9 h). Outcomes were coded 1 = “Yes”, 0 = “No”. All analyses used SI-consistent units (hours; kg/m² for body mass index).

### 2.5 Statistical analysis

Multivariable logistic regression modelled each outcome as a function of sleep group with adequate sleep as the reference, adjusting for age (category midpoint), sex, body mass index, smoking status (never/former/current), alcohol use, physical activity, and self-rated general health (5-level ordinal). Complete-case analysis yielded n=213,489 for 2022 and n=159,125 for 2020. Effect estimates are reported as adjusted odds ratios with 95% CIs; continuous exposures are summarised as medians with interquartile ranges (IQR), and network-density uncertainty as bootstrap standard deviations and 95% CIs. Two-sided *P*<0.05 was considered significant.

To verify the U shape rather than assume it, we fitted a second model containing centred linear (SleepHours − 7) and quadratic terms; a positive, significant quadratic coefficient establishes convexity, and the fitted nadir was computed as 7 − β₁/(2β₂). Wald tests compared the long- and short-sleep coefficients directly. For the pandemic analysis, separate 2020 and 2022 models were fitted, and a sleep-group × COVID-19-positivity product term was tested by a joint Wald test (2 df) in 2022. No correction for multiple testing was applied; *P* values should be interpreted accordingly. All analyses were performed in Python 3.13 (pandas 3.1, NumPy 2.5, statsmodels 0.15, SciPy 1.18, Matplotlib 3.11, NetworkX 3.7).

#### Survey-weighted sensitivity analysis

Weighted point estimates were obtained by pseudo-maximum-likelihood logistic regression with _LLCPWT normalised to mean 1. Variances were estimated by Taylor linearisation for the stratified, single-PSU-per-record design: with score contributions ui = wixi(yi − pi) and weighted information J, Cov(β^) = J⁻¹[Σh nh/(nh−1) Σi∈h(ui − ūh)(ui − ūh)′]J⁻¹. Strata containing a single PSU were centred on the grand mean of PSU totals, corresponding to survey.lonely.psu = “adjust”. Design degrees of freedom (number of PSUs minus number of strata) exceeded 300,000, so normal quantiles were used. We report the design effect (design-based SE² divided by naïve model-based SE²) and the Kish effective sample size (Σw)²/Σw². Design-weighted prevalences and domain shares used the same linearisation applied to ratio estimators, and weighted φ coefficients replaced simple correlations in the network analysis. This reproduces the estimator used by R survey::svyglm and is equivalent to the CDC-recommended SAS specification WEIGHT _LLCPWT; STRATA _STSTR; CLUSTER _PSU. Each survey wave was analysed separately, so no cross-year weight pooling was required.

### 2.6 Comorbidity-network analysis

Within each 2022 sleep stratum we built a network whose nodes were the 10 conditions and whose edges were pairwise **φ coefficients** — equivalently, Pearson correlations between two binary variables — computed as a 10 × 10 correlation matrix in float64 arithmetic. We defined **network density** as mean |φ| over all 45 unique edges, **node strength** as the sum of |φ| across a node’s edges, and the **Sleep Network Modulation Index (SSNM)** as density(stratum) − density(adequate). Uncertainty in density was estimated by 200 bootstrap resamples (seed 42). Because φ is a *marginal* association, we repeated the density comparison within strata defined by age (<65 versus ≥65 years), sex, and race/ethnicity.

### 2.7 Sensitivity, sex-specific, and race/ethnicity-specific analyses

For heart attack we re-estimated the sleep-group aORs (i) without self-rated general health, which may lie on the causal pathway or act as a collider; (ii) without the 4–12 hour range restriction; and (iii) under a tighter 5–11 hour restriction. To meet the AHA requirement for sex- and race/ethnicity-specific reporting, we additionally refitted the heart-attack and stroke models and recomputed network density within male and female strata and within four race/ethnicity strata (White non-Hispanic, Black non-Hispanic, Hispanic, and other/multiracial, the latter combining “Other race only” and “Multiracial” because of small numbers). These subgroup analyses are exploratory, unweighted, and not corrected for multiplicity. Separately, a design-weighted sensitivity analysis replicated all principal models on the official CDC LLCP 2020 and 2022 microdata (Section 2.5, Results 3.8, Table S2, Figure S1).

### 2.8 Data availability

In accordance with the AHA Journals’ implementation of the Transparency and Openness Promotion (TOP) Guidelines, the data, analytic methods, and study materials that support the findings of this study are available to other researchers for purposes of reproducing the results or replicating the procedure, subject to the data-use terms of the source repositories. All source data are publicly accessible: the 2020 and 2022 Behavioral Risk Factor Surveillance System public-use microdata, codebooks, and complex-design variables (_LLCPWT, _STSTR, _PSU) are distributed without charge by the US Centers for Disease Control and Prevention (https://www.cdc.gov/brfss/annual_data/annual_data.htm); Sleep Heart Health Study polysomnographic annotations are distributed by the National Sleep Research Resource (https://sleepdata.org/) to qualified investigators under its data-use agreement, which does not permit redistribution of the source EDF/XML files. The subject-level polysomnographic phenotype table derived for this analysis (shhs_features.csv) is supplied as Supplemental Material so that the objective-axis results can be reproduced without re-accessing the restricted source files.

The cleaned derivative CSV files used for the primary analyses, all derived analytic tables (phi_*.csv, prevalence_by_sleep.csv, prev_by_sleep_2022.csv), all result files (*.json, including survey_weighted_results.json), the supplementary tables (Tables S1 and S2), and all analysis and plotting scripts (*.py, including survey_design.py and survey_weighted_analysis.py) accompany this submission as Supplemental Material (Data S1 and Code S1); additional details about the analytical methods are available from the corresponding author on reasonable request. No new individual-level participant data were generated in this secondary analysis, and no data were obtained under an agreement restricting disclosure other than the NSRR terms stated above.

## Results

### 3.1 Study populations

Of 242,392 respondents with plausible 2022 sleep durations, 32.4% reported short sleep, 64.3% adequate sleep, and 3.2% long sleep. The SHHS PSG archive contributed 8,444 scored full-night recordings.

### 3.2 Axis 1: objective sleep is a continuous, severely distributed, and architecturally disrupted phenotype

The PSG-derived axis was strongly right-skewed (Figure 2A): median AHI 31.8 events/h (IQR, 19.0– 51.0; mean, 37.9; maximum, 286.9). Severity bands were 1.4% normal, 14.7% mild, 30.6% moderate, and 53.2% severe. Sleep architecture was correspondingly fragmented (Figure 2B): median sleep efficiency 69% (IQR, approximately 60–77%), with 93% of recordings below the 85% clinical threshold; median TST 6.13 hours against a TIB of 8.78 hours; slow-wave (N3) sleep 16% and REM 20% of TST (Figure 2B expresses the same stages relative to TIB). AHI and arousal index were moderately correlated (Spearman ρ=0.46; Figure 2C), consistent with respiratory events driving cortical micro-arousals. **Interpretive caveat.** SHHS was explicitly assembled to study SDB and is therefore enriched for it; NSRR event scoring may also be more sensitive than routine clinical scoring. The absolute AHI prevalences above are **cohort-specific and must not be read as population estimates**. What transfers to the model is structural: objective sleep is a graded, measurable pathophysiological axis with substantial within-cohort variance, largely invisible to a duration question.

**Figure 2.**
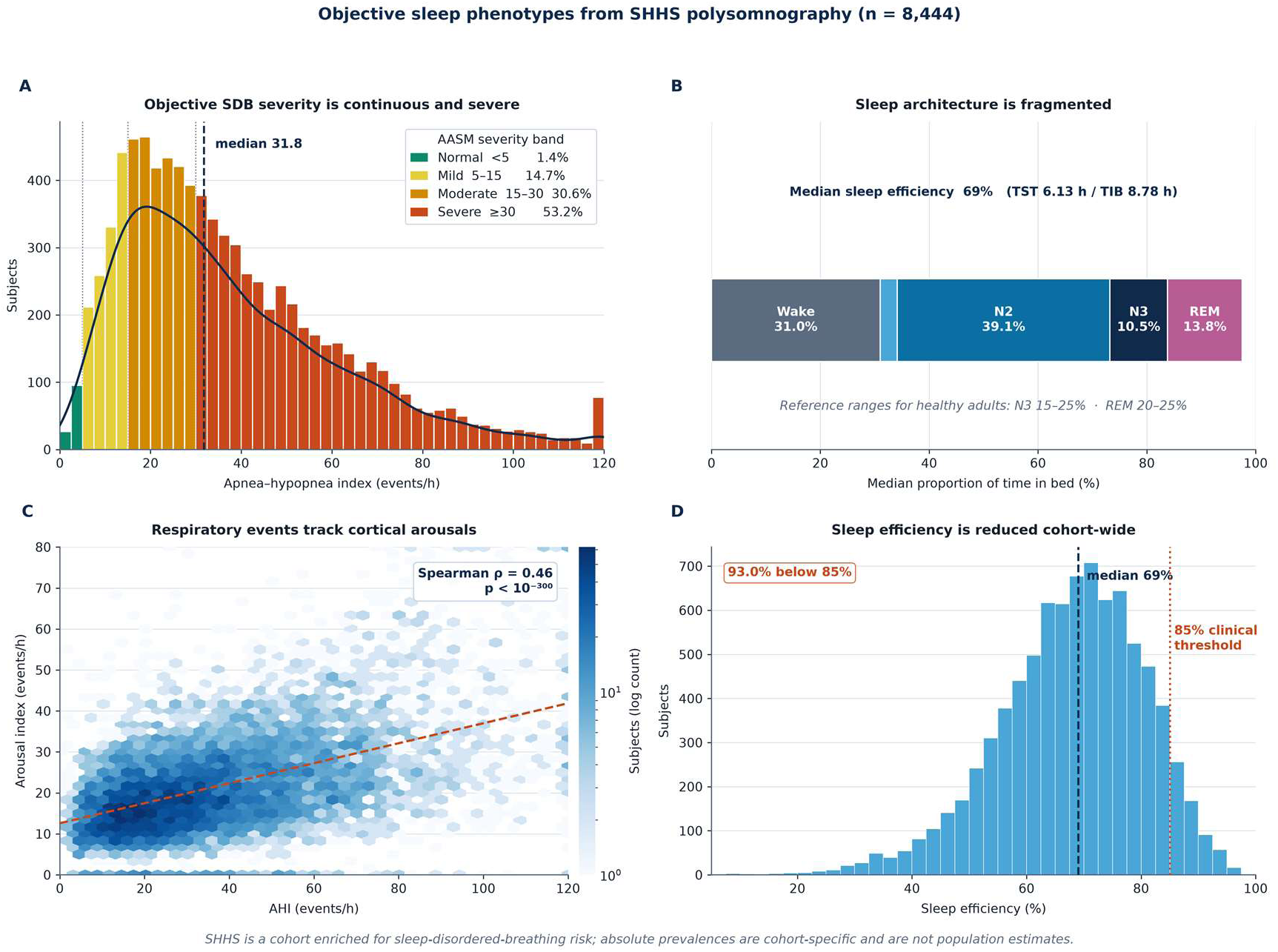
Objective sleep phenotypes in SHHS (n=8,444). (A) AHI distribution coloured by conventional severity band, with kernel-density overlay and median. (B) Median sleep architecture as a proportion of time in bed. (C) Hexbin density of AHI versus arousal index with linear fit and Spearman ρ. (D) Sleep-efficiency distribution with the 85% clinical threshold. SHHS is an SDB-enriched cohort; prevalences are not population estimates.

### 3.3 Axis 2: a formally verified U shape, with primary risk nadirs inside the recommended range

Crude prevalence rose from adequate to long sleep for nearly every condition (heart attack 4.76% → 6.04% → 11.27%; stroke 3.58% → 4.43% → 9.43%). After full adjustment, both short and long sleep remained independently associated with higher risk for 8 of the 9 modelled outcomes; the sole exception was long sleep and arthritis (aOR, 0.97; 95% CI, 0.92–1.03) (Table 1, Figure 4).

**Table 1.** Adjusted odds ratios (95% CI) by sleep duration; reference = adequate sleep (7–9 hours). BRFSS 2022, n=213,489.

| **Outcome** | **Short sleep (<7 h)** | **Long sleep (>9 h)** |
|---|---|---|
| Heart attack | 1.24 (1.19–1.30) | 1.30 (1.19–1.42) |
| Angina | 1.13 (1.09–1.18) | 1.12 (1.02–1.22) |
| Stroke | 1.12 (1.06–1.17) | 1.42 (1.29–1.57) |
| Diabetes | 1.10 (1.07–1.14) | 1.27 (1.19–1.36) |
| Kidney disease | 1.10 (1.05–1.15) | 1.26 (1.15–1.38) |
| Chronic obstructive pulmonary disease | 1.27 (1.22–1.32) | 1.28 (1.17–1.39) |
| Asthma | 1.17 (1.14–1.20) | 1.13 (1.05–1.21) |
| Depressive disorder | 1.19 (1.16–1.22) | 1.82 (1.72–1.94) |
| Arthritis | 1.27 (1.24–1.30) | 0.97 (0.92–1.03) |

*Models adjusted for age, sex, body mass index, smoking, alcohol use, physical activity, and general health. Nine conditions were modelled as regression outcomes; skin cancer was retained as a network node only. All estimates except long-sleep arthritis were significant at P<0.05.*

The U shape was confirmed formally: the quadratic term was positive and highly significant for heart attack (β₂=0.036, *P*=1.6×10⁻³²), stroke (0.038, *P*=3.4×10⁻³²), depressive disorder (0.057, *P*=3.7×10⁻¹⁹³), and diabetes (0.020, *P*=6.3×10⁻²⁰). Fitted nadirs were 7.85, 7.28, 7.26, and 7.38 hours respectively — all within the 7–9 hour range recommended for adults (Figure 3). The design-weighted replication preserved convexity for all four outcomes but widened the fitted nadir range to 6.77–8.20 hours, indicating that curvature is robust whereas the precise nadir is design-sensitive (Section 3.8).

**Figure 3.**
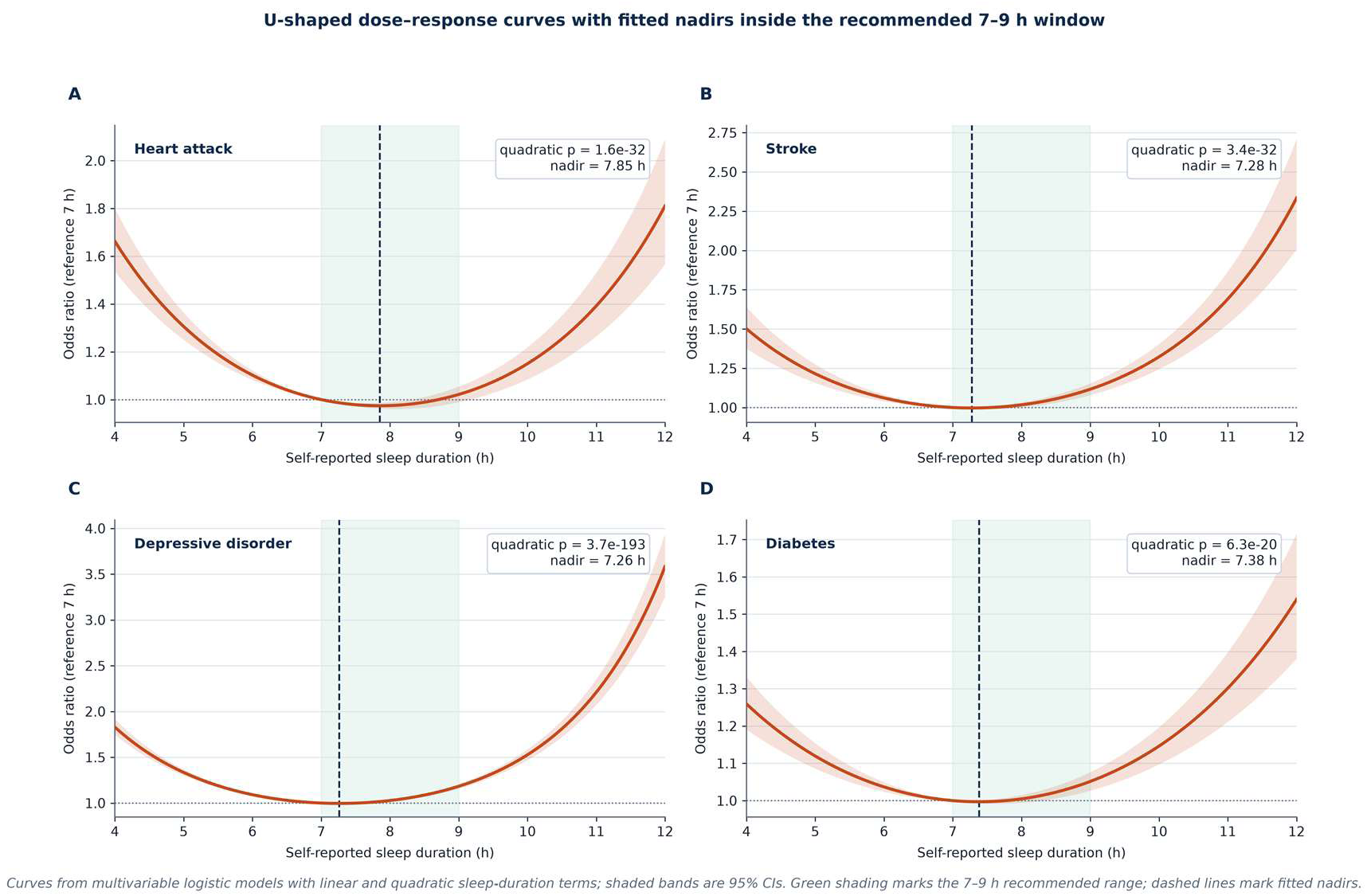
Fitted U-shaped dose–response curves (reference 7 hours) with 95% confidence bands for heart attack, stroke, depressive disorder, and diabetes. Green shading marks the recommended 7–9 hour range; dashed vertical lines mark fitted nadirs. Models include centred linear and quadratic duration terms with full covariate adjustment. Design-weighted nadirs were 6.77–8.20 hours (Section 3.8).

**Figure 4.**
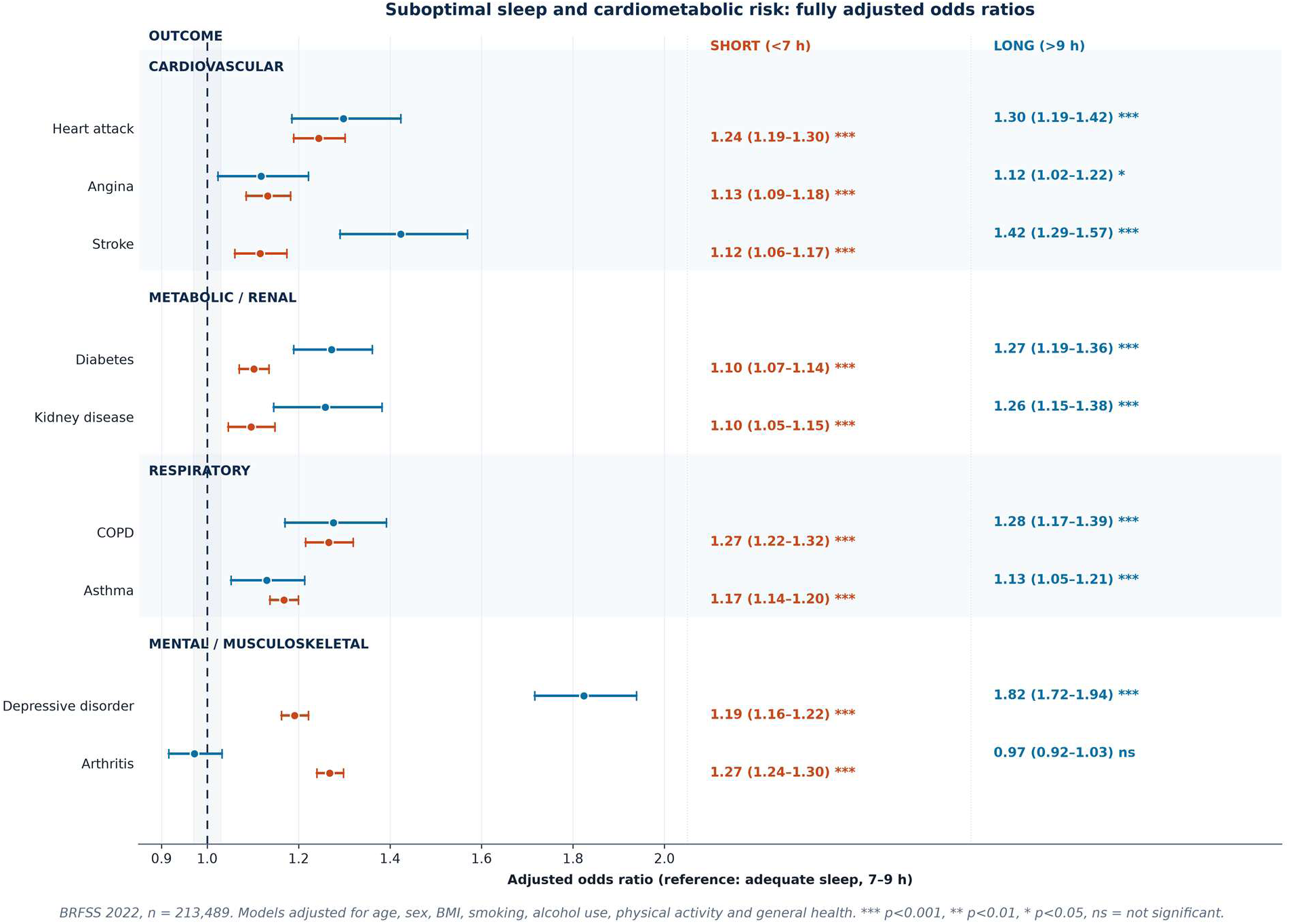
Forest plot of fully adjusted odds ratios for short (<7 hours, vermillion) and long (>9 hours, blue) sleep versus adequate sleep (7–9 hours) across nine outcomes, grouped by disease category. BRFSS 2022, n=213,489. Asterisks denote Wald *P*-value thresholds (**P*<0.001;*** P*<0.01; P*<0.05; ns, not significant).

The long-sleep arm was steeper than the short-sleep arm for stroke (difference in log-OR, 0.243; *P*=2.6×10⁻⁶) and depressive disorder (0.426; *P*=2.3×10⁻⁴¹), but the two arms did not differ for heart attack (0.043; *P*=0.37). Long sleep should therefore be read less as a direct causal insult than as a marker of underlying multimorbidity, frailty, or reverse causation.

### 3.4 Suboptimal sleep acts as a comorbidity-network amplifier

Treating the 10 conditions as nodes, the comorbidity network differed systematically across sleep strata (Figure 5A–C). Network density rose from **0.089** under adequate sleep to **0.112** under short sleep (+26.0%) and **0.108** under long sleep (+21.9%); the SSNM was +0.0231 and +0.0195, respectively (Table 2). Bootstrap 95% CIs did not overlap between the adequate and either suboptimal stratum, and the ordering was preserved in every demographic stratum examined: adults <65 years (+32.3% / +56.8%), adults ≥65 years (+27.5% / +19.3%), men (+24.8% / +23.0%), and women (+25.7% / +23.3%).

**Figure 5.**
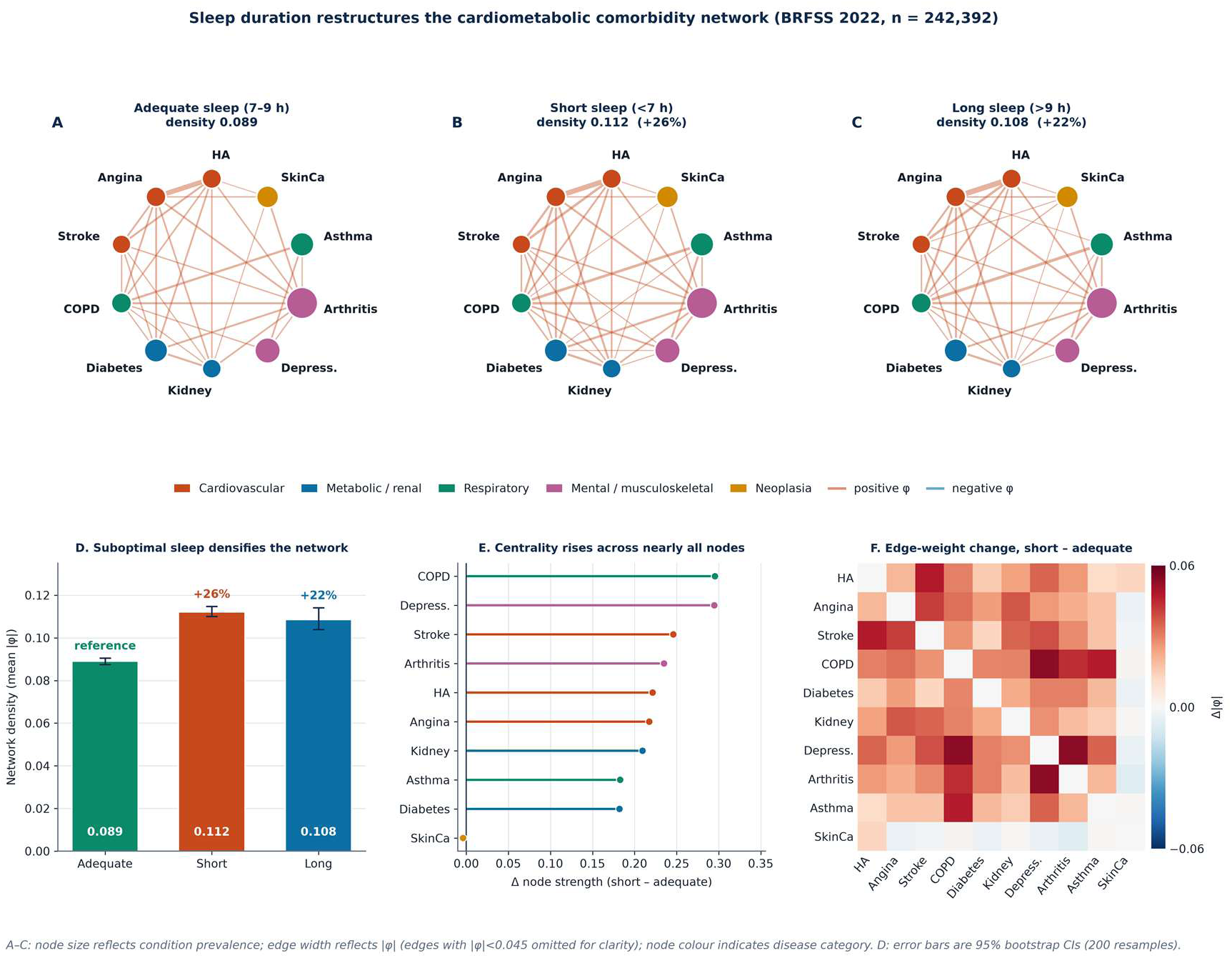
Sleep-stratified comorbidity networks (BRFSS 2022, n=242,392). (A–C) Networks under adequate, short, and long sleep; node size reflects condition prevalence, node colour disease category, edge width |φ| (edges with |φ|<0.045 omitted for clarity). (D) Network density with 95% bootstrap CIs (200 resamples). (E) Change in node strength, short minus adequate. (F) Heatmap of change in |φ|, short minus adequate.

**Table 2.**
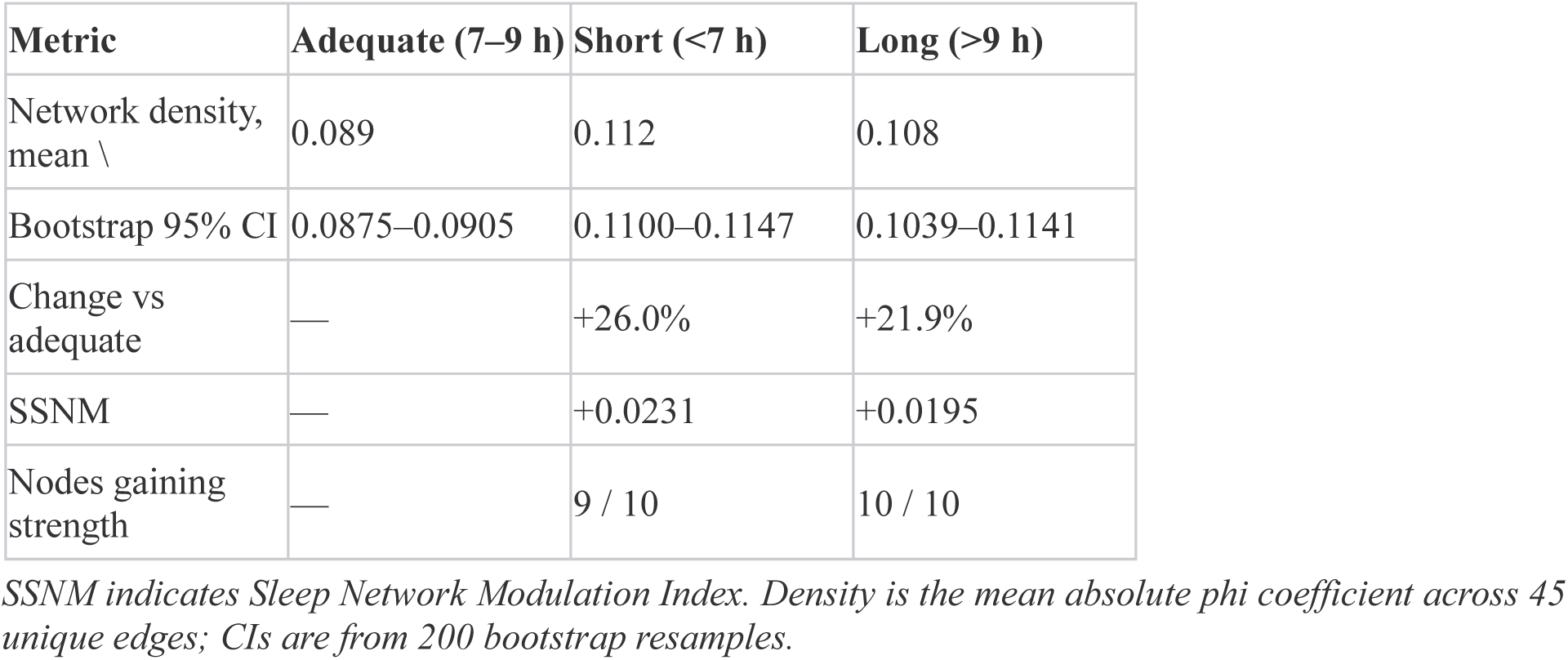
Comorbidity-network metrics by sleep stratum (BRFSS 2022, n=242,392).

| Metric | Adequate (7–9 h) | Short (<7 h) | Long (>9 h) |
|---|---|---|---|
| Network density, mean \ | 0.089 | 0.112 | 0.108 |
| Bootstrap 95% CI | 0.0875–0.0905 | 0.1100–0.1147 | 0.1039–0.1141 |
| Change vs adequate | — | +26.0% | +21.9% |
| SSNM | — | +0.0231 | +0.0195 |
| Nodes gaining strength | — | 9 / 10 | 10 / 10 |

*SSNM indicates Sleep Network Modulation Index. Density is the mean absolute phi coefficient across 45 unique edges; CIs are from 200 bootstrap resamples.*

At node level, 9 of 10 conditions gained strength under short sleep (skin cancer was essentially unchanged, Δ=−0.004), with the largest increases for chronic obstructive pulmonary disease (+0.296), depression (+0.295), stroke (+0.246), and arthritis (+0.235); under long sleep all 10 gained strength (range, +0.054 to +0.243). At edge level the change was broadly distributed rather than driven by a single pathway: existing associations strengthened, including chronic obstructive pulmonary disease–depression (φ 0.085 → 0.139), depression–arthritis (0.088 → 0.142), heart attack–stroke (0.152 → 0.199), and chronic obstructive pulmonary disease–asthma (0.172 → 0.219). Suboptimal sleep therefore **amplifies coupling across the network** rather than creating entirely new disease pairs — a distinction we emphasise because a naive reading of sparse marginal correlations can suggest spurious “new” edges. A design-weighted, nine-condition network reproduced the same ordering (Section 3.8).

### 3.5 Sex- and race/ethnicity-specific analyses

The direction of the network effect was concordant in every sex and race/ethnicity stratum (Table S1). Network density rose from adequate to short sleep by 23.0% in men and 23.5% in women, and by 23.9%, 18.0%, 17.4%, and 13.3% in White non-Hispanic, Black non-Hispanic, Hispanic, and other/multiracial respondents, respectively; long sleep exceeded adequate sleep in all six strata. The heart-attack aORs for short sleep were 1.18 (95% CI, 1.11–1.25) in men and 1.35 (1.26–1.46) in women, suggesting a stronger association in women, although no formal sex-interaction test was prespecified. Point estimates were less precise in the smaller race/ethnicity strata, and several CIs crossed 1.0; these analyses are exploratory and should not be read as evidence of heterogeneity.

### 3.6 Exploratory era and COVID-19 contrasts

Comparing the pre-pandemic 2020 wave with the 2022 wave, the sleep–heart-attack association strengthened for both short (aOR, 1.17; 95% CI, 1.12–1.22 → 1.24, 1.19–1.30) and long sleep (1.11, 1.01–1.21 → 1.30, 1.19–1.42), whereas the sleep–stroke association was essentially stable for long sleep (1.41 → 1.42) and slightly attenuated for short sleep (1.13 → 1.12) (Figure 6A, Table 3). Because the 2020 HeartDisease item is a composite question while 2022 records HadHeartAttack, this comparison is directional rather than a like-for-like replication.

**Figure 6.**
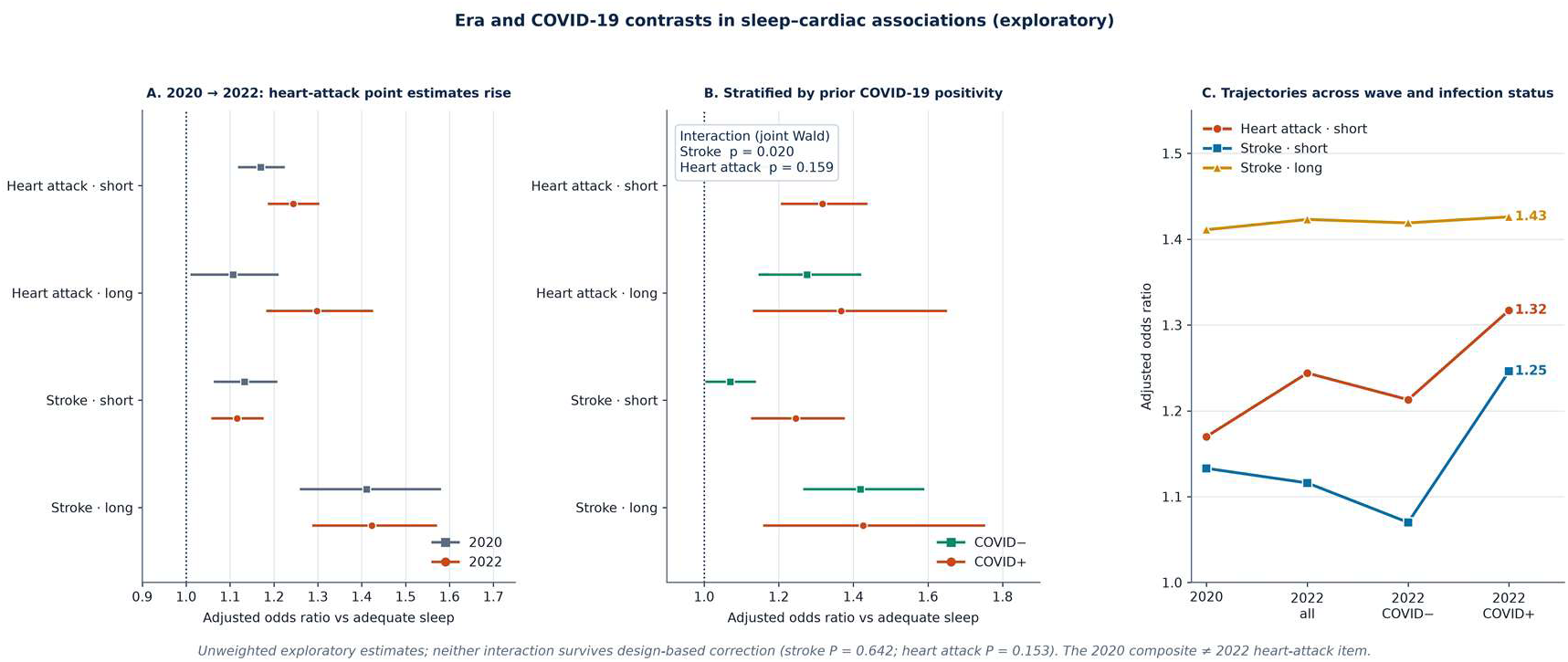
Era and COVID-19 analyses. (A) Adjusted odds ratios in 2020 versus 2022. (B) 2022 estimates stratified by prior COVID-19 positivity, with joint Wald interaction *P* values. (C) Amplification trajectories across survey wave and infection status. Error bars are 95% CIs. The 2020 composite heart-disease item and 2022 heart-attack item are not identical constructs. All estimates and interaction *P* values are unweighted and exploratory; under design-based variance estimation neither interaction was significant (Section 3.8).

**Table 3.** Era comparison and COVID-19-stratified adjusted odds ratios (reference = adequate sleep).

| **Outcome / sleep group** | **2020 aOR (95% CI)** | **2022 aOR (95% CI)** | **2022 COVID-negative** | **2022 COVID-positive** | **Interaction *P*** |
|---|---|---|---|---|---|
| Heart attack / short | 1.17 (1.12–1.22) | 1.24 (1.19–1.30) | 1.21 (1.15–1.28) | 1.32 (1.21–1.44) | 0.159 |
| Heart attack / long | 1.11 (1.01–1.21) | 1.30 (1.19–1.42) | 1.28 (1.15–1.42) | 1.37 (1.13–1.65) | 0.159 |
| Stroke / short | 1.13 (1.07–1.21) | 1.12 (1.06–1.17) | 1.07 (1.01–1.14) | 1.25 (1.13–1.37) | 0.020 |
| Stroke / long | 1.41 (1.26–1.58) | 1.42 (1.29–1.57) | 1.42 (1.27–1.59) | 1.43 (1.16–1.75) | 0.020 |

*aOR indicates adjusted odds ratio; COVID-19, coronavirus disease 2019. The 2020* HeartDisease *item is a composite and is not identical to the 2022* HadHeartAttack *item.*

Stratifying 2022 by prior COVID-19 positivity showed a graded increase in point estimates (Figure 6B): for short sleep, the heart-attack aOR rose from 1.21 (COVID-negative) to 1.32 (COVID-positive), and the stroke aOR from 1.07 to 1.25. The formal **sleep × infection interaction**, however, reached significance only for stroke (χ²=7.83, 2 df, *P*=0.020) and not for heart attack (χ²=3.68, 2 df, *P*=0.159). We therefore described the COVID-19 finding as *outcome-specific effect modification* rather than a general amplification. Under design-based variance estimation, however, neither interaction remained significant (stroke *P*=0.642; heart attack *P*=0.153; Section 3.8); the era and infection contrasts are consequently treated as exploratory and are not used to support the principal conclusions.

### 3.7 Sensitivity analyses

The heart-attack estimates were directionally robust across specifications (Table 4). Removing self-rated general health **strengthened** the associations (short, 1.45; long, 1.62), suggesting that general health may act as a mediator or collider rather than a pure confounder; the primary model is therefore conservative. Relaxing the range restriction (1.26 / 1.31) and tightening it to 5–11 hours (1.19 / 1.27) left the conclusions unchanged.

**Table 4.** Sensitivity of the heart-attack association to model specification.

| **Specification** | **n** | **Short sleep aOR** | **Long sleep aOR** |
|---|---|---|---|
| Primary model | 213,489 | 1.24 | 1.30 |
| Without general health | 213,489 | 1.45 | 1.62 |
| No 4–12 hour restriction | 216,264 | 1.26 | 1.31 |
| Restricted to 5–11 hours | 207,520 | 1.19 | 1.27 |

*aOR indicates adjusted odds ratio.*

### 3.8 Design-weighted sensitivity analysis

The official LLCP files yielded 324,335 (2020) and 344,010 (2022) complete-case records across 1,592 and 2,238 design strata. Mean final weights were 636 and 572, and Kish effective sample sizes were 56,745 and 75,927 — approximately one fifth of the nominal counts. Design-weighted 2022 sleep-duration shares were 35.3% (95% CI, 34.9–35.6%) short, 61.8% (61.5–62.2%) adequate, and 2.9% (2.8– 3.0%) long; design-weighted prevalences were 4.10% for heart attack, 3.29% for stroke, and 20.95% for depressive disorder. These are population estimates and are not interchangeable with the unweighted sample quantities reported above.

Across the 18 sleep-group coefficients estimated in 2022, the median design effect was 4.43 (range, 3.85– 5.21), with 2020 coefficients reaching 6.51: ignoring the survey design had understated standard errors by approximately a factor of two. Nevertheless, the principal cardiac associations retained both direction and significance. Design-weighted 2022 aORs were 1.30 (95% CI, 1.20–1.40) and 1.17 (1.02–1.34) for heart attack under short and long sleep; 1.15 (1.06–1.26) and 1.51 (1.30–1.77) for stroke; and 1.27 (1.22–1.32) and 1.86 (1.70–2.04) for depressive disorder (Table S2, Figure S1). The quadratic term remained positive and significant for all four outcomes tested (weighted *P*≤1.1×10⁻³), although fitted nadirs shifted to 6.77– 8.20 hours. The nine-condition comorbidity network again densified under suboptimal sleep (weighted mean |φ| 0.106 → 0.132 for short sleep, +24.5%; 0.134 for long sleep, +25.7%), compared with 0.100 → 0.129 and 0.118 under unweighted estimation on the same records.

Not all findings were design-robust. Short-sleep associations with diabetes (1.03; 95% CI, 0.98–1.09) and kidney disease (1.08; 0.99–1.17), and long-sleep associations with angina (1.01; 0.88–1.16), arthritis (1.01; 0.91–1.11), and asthma (1.11; 0.99–1.24), crossed 1.0 after design-based correction. Most importantly, the sleep × COVID-19-positivity interaction that was nominally significant for stroke in the unweighted analysis (χ²=7.83, *P*=0.020) was not significant under design-based variance estimation (χ²=0.89, *P*=0.642; heart attack χ²=3.76, *P*=0.153). We therefore regard era and infection contrasts as exploratory.

The unweighted replication on official microdata closely reproduced the main-text estimates (2022 short-sleep heart attack 1.24 versus 1.24; 2020 long-sleep stroke 1.41 versus 1.41; Table S2, Panel F), supporting the validity of the recoding. Because the derivative files carry no respondent identifier, weights could not be attached to the exact primary records; the weighted analysis is therefore an independent, same-specification replication rather than a reweighting of the primary dataset.

## Discussion

### 4.1 Principal findings

Three strands of evidence converge. First, objective PSG defines a continuous, graded, and frequently severe axis of sleep pathology that a duration question cannot access. Second, subjective duration carries its own, formally U-shaped association with cardiometabolic disease, with fitted nadirs falling inside the guideline range. Third — and most unexpectedly — suboptimal sleep is associated not merely with higher risk for individual diseases but with a **denser, more strongly coupled multimorbidity network**, an effect that survives bootstrap resampling and stratification by age, sex, and race/ethnicity. A design-weighted replication on official CDC microdata preserved all three strands despite a median design effect of 4.43. Superimposed era and infection contrasts were exploratory and did not survive design-based variance correction (Section 3.8).

### 4.2 Comparison with prior work

The U-shaped duration association reproduces a substantial literature^5–8^; our contribution here is to verify its curvature formally, locate the primary nadirs within the recommended 7–9 hour window (while showing that the precise nadir is design-sensitive; Section 3.8), and show that the two arms are not symmetric — long sleep was significantly steeper for stroke and depression. That asymmetry accords with the view that prolonged sleep often reflects illness burden rather than causing it.^5^ The SDB findings are consistent with the original SHHS reports and the contemporary AHA scientific statement,^1–4^ and with the well-documented underdiagnosis of obstructive sleep apnoea^11^; we extend them by placing SDB on a separate axis from self-reported duration rather than treating the two as interchangeable sleep exposures. A multidimensional view of sleep health^9^ and recent evidence linking habitual duration to cardiovascular health and utilisation^10^ provide the conceptual backdrop for that separation.

Network medicine has mapped disease co-occurrence at population scale and shown that the measurement of co-occurrence shapes the inferred network.^12–15^ To our knowledge, however, sleep duration has not previously been examined as a *modulator* of that network. The SSNM offers a compact, reproducible summary that could be tracked longitudinally or compared across cohorts.

### 4.3 Mechanistic interpretation

A plausible common substrate is that adequate sleep acts as a systemic homeostatic reset. Short or fragmented sleep engages sympathetic activation, hypothalamic–pituitary–adrenal dysregulation, impaired glycaemic control, and low-grade inflammation^17,18^; intermittent hypoxia from SDB adds oxidative stress and endothelial dysfunction.^1–4^ If such processes raise the background susceptibility of many organ systems at once, previously weak disease–disease associations will strengthen in parallel — exactly the broad, distributed increase in edge weights we observed. The nominally stronger estimates among COVID-positive respondents are compatible with persistent post-viral autonomic and inflammatory sequelae compounding sleep-related risk,^19^ but this interaction did not survive design-based variance correction and should not be read as established effect modification.

### 4.4 Clinical and public-health implications

The Two-Axis model has a direct screening corollary: a reassuring self-report of 7–9 hours does **not** exclude severe SDB. In a patient with hypertension, atrial fibrillation, stroke, or resistant metabolic risk, objective sleep assessment should be considered irrespective of reported duration. Conversely, the steep long-sleep arm argues that a report of >9 hours deserves clinical attention in its own right as a marker of occult multimorbidity rather than being dismissed as healthy behaviour. The American Heart Association’s Life’s Essential 8 framework now explicitly includes sleep among the components of cardiovascular health,^20^ and our findings suggest that this component should be measured on both axes rather than reduced to a single duration item. At population level, sleep is a modifiable exposure sitting upstream of several simultaneously coupled disease clusters, which may make it a comparatively efficient prevention target.

### 4.5 Strengths and limitations

Strengths include the sample size, the use of both gold-standard PSG and a large national health survey, formal curvature testing, bootstrap and stratified sensitivity analyses for the network result, and conservative adjustment.

Several limitations temper interpretation. **(i) Cohort non-overlap.** SHHS and BRFSS are independent populations, so we cannot estimate the within-person correlation between objective SDB and subjective duration; separability of the axes is a theoretically motivated, separately supported proposition, not a directly tested individual-level correlation. A definitive test requires a cohort with linked PSG and survey data. **(ii) Cross-sectional design.** Directionality cannot be inferred, and reverse causation is especially plausible for long sleep. **(iii) SHHS enrichment.** Absolute AHI prevalences reflect deliberate oversampling of SDB risk and NSRR scoring conventions; they are not generalisable. **(iv) Self-report.** A single-item sleep-duration question is subject to misclassification, and outcomes are self-reported physician diagnoses; shared-method bias cannot be excluded. Range restriction mitigates but does not eliminate the former. **(v) Residual confounding.** Socioeconomic position, medication, sleep apnoea treatment, and comorbidity severity were unavailable. **(vi) Marginal network edges.** φ coefficients are unadjusted; although age, sex, and race/ethnicity stratification reproduced the pattern, other shared determinants could contribute, and the choice of co-occurrence measure may affect absolute densities. **(vii) Outcome harmonisation.** The 2020 composite HeartDisease item and 2022 HadHeartAttack are not identical constructs. **(viii) Multiple testing.** No multiplicity correction was applied; the very small *P* values for the U-shape and arm-contrast tests are robust to any reasonable correction, whereas the stroke interaction (*P*=0.020) and the exploratory subgroup comparisons should be regarded as suggestive; the stroke interaction was in fact not significant after design-based variance correction (Section 3.8). **(ix) Adjustment set.** Self-rated general health may be a mediator or collider; its inclusion attenuates estimates, so reported aORs are conservative.

### 4.6 Future directions

Priority next steps are validation in cohorts with jointly measured PSG and self-reported sleep (for example the Multi-Ethnic Study of Atherosclerosis or linked SHHS follow-up), confirmation of the design-weighted estimates with an independent survey implementation (R survey or SAS SUDAAN), prospective testing of whether the SSNM predicts incident multimorbidity, and trials examining whether treating SDB benefits the *silent SDB* quadrant differently from the *behavioural risk* quadrant.

## Conclusions

Sleep and cardiac risk are better described by **two dissociable axes** than by a single number. Objective sleep-disordered breathing and subjective sleep duration are each independently prognostic, and suboptimal sleep further acts as an amplifier of the cardiometabolic comorbidity network — a structural effect that single-outcome analyses cannot reveal. The principal associations were preserved in a design-weighted replication on official CDC microdata, whereas pandemic-era and infection-related contrasts were exploratory and did not survive design-based correction. Assessing sleep on both axes, rather than asking only about hours, may uncover a large and currently silent high-risk population.

## Acknowledgments

**Author contributions.** Q.D., N.M., T.A. and D.P. conceived and designed the study. Q.D. and N.M. curated the data and performed the statistical analyses, including the design-weighted replication. T.A. prepared the figures and the Supplemental Material. D.P. acquired funding and supervised the project. Dr Pang had full access to all the data in the study and takes responsibility for its integrity and the data analysis. All authors read and approved the final manuscript. All persons named have agreed to this acknowledgement.

**Non-author acknowledgements.** Not applicable.

**Generative-AI disclosure.** No artificial-intelligence tool qualifies for or is listed as an author, and the authors take full responsibility for the accuracy, validity, and originality of the content. No data within any figure were enhanced, obscured, moved, removed, or introduced by digital manipulation or generative-AI image technology.

## Sources of Funding

This work was supported by the Clinical Cultivation Innovative Team for Acute Large Vessel Occlusion Stroke (grant number 2026-XEQ-01).

## Disclosures

None. The authors declare that they have no competing interests.

## Non-standard Abbreviations and Acronyms

AHI: apnea–hypopnea index
aOR: adjusted odds ratio
BRFSS: Behavioral Risk Factor Surveillance System
CDC: Centers for Disease Control and Prevention
DE: design effect
LLCP: landline and cellular telephone survey data file
NSRR: National Sleep Research Resource
PSG: polysomnography
PSU: primary sampling unit
SDB: sleep-disordered breathing
SHHS: Sleep Heart Health Study;
SSNM: Sleep Network Modulation Index
TIB: time in bed
TST: total sleep time
φ: phi coefficient.

## Supplemental Material

- **Table S1.** Sex- and race/ethnicity-stratified sensitivity analyses (Table_S1_demographic_strata.md).
- **Table S2.** Survey-weighted sensitivity analysis on official CDC BRFSS LLCP 2020/2022 microdata (Table_S2_weighted_sensitivity.md).
- **Data S1.** Derived analytic datasets (shhs_features.csv, phi_*.csv, prevalence_by_sleep.csv, prev_by_sleep_2022.csv) and all result files (*.json, including survey_weighted_results.json).
- **Code S1.** All analysis and plotting scripts (*.py, including survey_design.py, survey_weighted_analysis.py, brfss_probe.py, make_weighted_figure.py).
- **Checklist S1.** Completed STROBE cross-sectional checklist.
- **Report S1.** Technical report on the design-weighted replication (survey_weighted_sensitivity.md; internal, not for upload).

**Figure S1.**
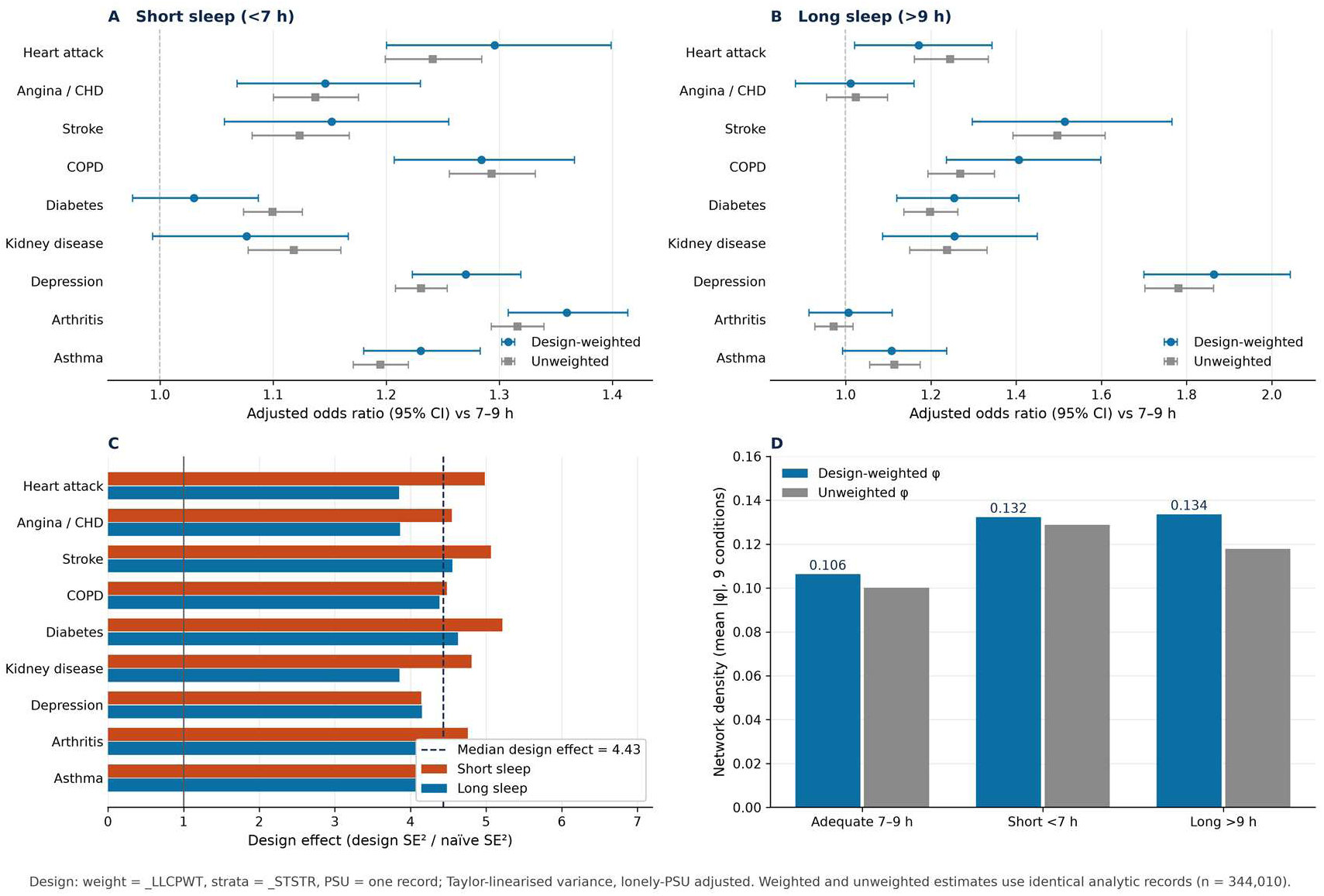
Design-weighted versus unweighted adjusted odds ratios, design effects, and network density (figures_jaha/figS1_weighted_sensitivity.pdf). Survey-weighted sensitivity analysis on CDC BRFSS LLCP 2022 microdata

## Notes

### Competing Interest Statement

The authors have declared no competing interest.

### Clinical Trial

This is not a clinical trial

